# Connectivity between long-term care homes and subsequent SARS-CoV-2 outbreaks

**DOI:** 10.1101/2025.01.08.25320217

**Authors:** Yiqing Xia, Huiting Ma, Kamil Malikov, Sharon E. Straus, Christine Fahim, Gary Moloney, Qing Huang, Sahar Asgari, Jamie M. Boyd, Irene Ferro, Jaimie Johns, Kamran Khan, Jaydeep Mistry, Linwei Wang, Adrienne K. Chan, Stefan D. Baral, Mathieu Maheu-Giroux, Sharmistha Mishra

**Author notes:** Corresponding Author: Sharmistha Mishra, Mailing address: Room 315, MAP Centre for Urban Health Solutions, Li Ka Shing Knowledge Institute, University of Toronto, 209 Victoria St., Toronto, ON Canada M5B 1T8.

## Abstract

**Objectives:** To describe the relationship between individual workers employed at more than one LTCH (inter-LTCH connectivity) across long-term care homes (LTCH) and SARS-CoV-2 outbreaks.

**Design:** A retrospective cohort study using long-term care home surveillance and mobile geolocation data.

**Setting:** Using data observed between February 26^th^, 2020, and August 31^st^, 2020, from Ontario, the province where close to one-third of the Canada’s SARS-CoV-2 cases among long-term care homes residents were reported.

**Participants:** We included all 179 LTCH in the Greater Toronto Area (population 6.7 million, where close to 50% of Ontario population resides).

**Exposures:** The main exposure of interest was the inter-LTCH connectivity, generated from geographic position system location data procured across apps on different platforms.

**Main outcomes and measures:** Three outcomes were examined: 1) at least one SARS-CoV-2 diagnosis among residents, 2) cumulative cases among residents in each facility, and 3) time to first outbreak.

**Results:** The median degree of connectivity for LTCH that experienced an outbreak (59%; 106/179) was 1.2 times the degree of those without an outbreak (6 compared to 5). LTCH with higher inter-LTCH connectivity also had larger numbers of residents and beds, and were more likely to have for-profit ownership. After adjusting for facility-level and neighbourhood-level factors, every additional connection to another LTCH increased the odds of an outbreak in the respective LTCH by 8% (adjusted odds ratio=1.08, 90% credible interval [CrI]: 1.02-1.09). Inter-LTCH connectivity was also associated with higher risk of earlier occurrence of a first SARS-CoV-2 case (adjusted hazard ratio=1.05, 90%CrI: 1.02-1.09), but not with outbreak size.

**Conclusions and Relevance:** Staff cohorting was associated with reduced importation risk of SARS-CoV-2 cases into LTCH. However, findings suggest that once importation has occurred, other facility-level factors including facility infrastructure and staff benefits are more important in shaping outbreak size. Implementing these structural strategies to meet the LTCH workers and residents’ needs are pivotal to prevent and manage future respiratory virus outbreaks.

**Key points:** *Question:* Were movement of long-term care homes (LTCH) workers between facilities (staff connectivity) associated with the risk, size, and timing of SARS-CoV-2 outbreaks in these facilities during the first wave of the COVID-19 pandemic.

*Finding:* After adjusting for facility-level and neighbourhood-level factors, a higher degree of staff connectivity between LTCH was associated with a greater risk of outbreaks (2.2-fold the risk of a LTCH connected with 10 more other LTCHs) and a higher risk of experiencing an earlier outbreak (1.7-fold the hazard with 10 more staff connections with other LTCH). However, we did not observe an association between connectivity and the size of outbreaks.

*Meaning:* “One-site” strategy to cohort staff by facility and minimizing movement may reduce risk of pathogen importation. However, structural strategies (e.g. improve facility design and infrastructure) to reduce nosocomial transmission within these facilities remain pivotal to prevent and manage future respiratory virus outbreaks.

## Introduction

Long-term care home (LTCH) residents and workers were disproportionately affected by the COVID-19 pandemic (1). In Canada, LTCH residents accounted for 80% of COVID-19 related deaths during the first wave of the epidemic (2, 3). LTCH in Ontario, the most populous province of Canada, were among the hardest hit (4–6). During the first wave of the SARS-CoV-2 epidemic in early 2020, LTCH residents in Ontario accounted for nearly one-third of the total number of SARS-CoV-2 diagnoses and deaths among LTCH residents in Canada (7). In comparison, Ontario represents approximately 20% of Canada’s total number of LTCH residents (8, 9).

Even prior to the COVID-19 pandemic, respiratory virus outbreaks including influenza and SARS-CoV-1 have represented sustained threats to the health of residents in LTCH (10). The reasons for elevated risks of nosocomial transmissions in LTCH are multifactorial, including the vulnerability of the residents (e.g., old age, comorbidities), facilities that are not designed for infection control practices (e.g., communal settings and multiple-occupancy rooms), lacking protocols of infection control and personal protective equipment (11). Nosocomial transmissions refer to infections acquired among LTCH residents, in staff, or in visitors when pathogens are transmitted or acquired between persons within the facility. Thus, efforts to reduce nosocomial outbreaks largely centered on infection control practices (such as masking, hand hygiene, etc.), improved environmental cleaning and ventilation, and strategies to reduce the introduction of infections acquired outside the facility. The latter include infections acquired in the community or in other healthcare institutions if personnels work in multiple facilities. Thus, in addition to restrictions on visitors to LTCH (12), one of the earliest pandemic response strategies employed in Ontario was provincial guidance to restrict LTCH personnel from working in more than one congregate facility–strategy implemented across jurisdictions in Canada and other countries (4, 13, 14). That is, nurses, personal support workers, and other care providers had to choose one facility for employment, in an effort to reduce SARS-CoV-2 transmission between LTCHs.

However, movement of workers between different LTCH and other congregate settings (e.g. retirement homes, shelters) or home care, is common. This is especially true in the context of the shortage of healthcare workers across Canada (15). Working in multiple facilities is particularly common among part-time LTCH staff and agency-staff (i.e., temporary caregivers employed by a third party) (16). It is estimated that 24% of LTCH staff work in multiple facilities (17). The majority of LTCH staff who provide direct care for residents are personal support workers earning lower-than-average wages (18–20). In Ontario and across Canada, a confluence of exposure risks for SARS-CoV-2, outside healthcare facilities and within such facilities, led to disproportionate risks of infection among LTCH staff, especially but not limited to personal support workers (21). Chief among these risk factors for SARS-CoV-2 outbreaks were structural barriers to isolation if exposed or infected, such as lack of paid sick leave or crowded housing, as well as heightened risk of repeat exposures at workplaces and in the community (21, 22).

The rationale for restricting movement of staff to only work on one facility during pandemic was to limit the potential spread of SARS-CoV-2 between facilities (23). In the context of ongoing staffing shortages, however, there may have been negative consequences such as workloads exceeding capacity leading to burnout and moral injury among staff (24), and gaps in the quality of care (25, 26)–all of which might have contributed to the spread of SARS-CoV-2 (27). To date, it is unknown to what extent staff movement between long-term care facilities could explain outbreaks within congregated settings such as LTCH in Canada. The overarching goal of this study is to characterize the association between LTCH worker connectivity and SARS-CoV-2 outbreaks in the Greater Toronto Area (Canada). Specifically, we aim to examine whether the inter-facility connectivity was associated with 1) an outbreak in LTCH, 2) cumulative numbers of SARS-CoV2 cases in LTCH residents, and 3) time-to-first SARS-CoV-2 case in each LTCH.

## Methods

### Study design and study population

We conducted a retrospective cohort study of all 179 long-term care homes in the Greater Toronto Area, the largest urban center in Ontario, Canada from February 26^th^, 2020, to August 31^st^, 2020 (the first major wave of COVID-19 epidemic). The Greater Toronto Area includes Toronto, York, Peel, Halton, and Durham municipalities, where close to 50% of the Ontario’s population resides (6.7 million).

### Data sources

Aggregated facility-level total number of SARS-CoV-2 cases were extracted from the *Case and Contact Management System* (a central data repository for SARS-CoV-2 case and contact management (28)) and the *Integrated Public Health Information System* (a centralized information system for the reporting and surveillance of all Diseases of Public Health Significance (29)) in Ontario which includes all suspected and laboratory-confirmed cases of SARS-CoV-2 infections. Characteristics and occupancies of each LTCH were obtained from the *Client Profile* , a database containing information on LTCH attributes and residents information (30). Data were obtained from Ontario Ministry of Health and Long-Term Care under a data sharing agreement.

Aggregated LTCH facility-level mobility data were generated by BlueDot using anonymized geographic position system location data procured across apps on different platforms with users’ consent from Pelmorex and Veraset (31, 32). BlueDot summarized the mobility data over the study period at the level of each LTCH. Cumulative incidence of SARS-CoV-2 cases in the surrounding dissemination area (a small geographic unit with around 400-700 people; thereafter, “*neighborhood* ”) where the LTCH were located during study period was extracted from an online database that was verified by the Ontario Ministry of Health (33).

### Measures and Outcomes

The main exposure of interest was the movement of LTCH workers across facilities. Due to lack of administrative data, we used the overall degree of connectivity between facilities during the study period (thereafter, “*inter-LTCH connectivity* ”) as proxy. This is measured as the number of facilities a LTCH shares at least one smartphone connection with (34). A connection between two facilities was recorded when a mobile device makes a visit (a minimum of 30 minutes within the facility geographic unit) to one facility and makes another visit to a different facility within 14-days of visiting the first. We assumed the observed connectivity between facilities was due to worker movement, as the lockdown and visiting restriction measures were also implemented at the same time as the study period (35, 36), which limited the movement of the general population and banned visiting of LTCH residents.

We adjusted for facility-level factors that were related to COVID-19 outbreaks and transmission in LTCH, including for-profit ownership (Yes/No), total number of beds, proportion of private beds (1 bed per room), and the number of residents (37). Due to data availability limitations, we used the facility attributes on February 1^st^, 2020, and assumed they are constant throughout the study period given the short time frame of the study.

Additionally, we included an area-level measure that was associated with SARS-CoV-2 diagnosis in LTCH: number of cases in the surrounding community (23). Given the large heterogeneities in the numbers of cases within surrounding community, we categorized this measure into deciles: the LTCH was categorized as being located in a hotspot if the cumulative rate of SARS-CoV-2 diagnoses in the neighbourhood surrounding the LTCH during the study period was >300 per 100,000 population (33).

The primary outcome was the outbreak status (Yes/No) of a LTCH, which is defined as at least one SARS-CoV-2 case diagnosed among the residents (7). We examined two secondary outcomes: outbreak size (measured as the total number of SARS-CoV-2 cases in each LTCH during the study period); and time-to-first SARS-CoV-2 case since the beginning of the epidemic in Ontario (2020-02-26). A SARS-CoV-2 case was defined as a confirmed laboratory test using the polymerase chain reaction among residents (38).

### Statistical Analyses

To examine whether the inter-LTCH connectivity was associated with LTCH outbreaks, a Bayesian logistic regression model (Model 1) was built. Additionally, a Bayesian negative binomial model (Model 2) and a Bayesian Cox proportional hazard model (Model 3) were used to detect whether the inter-LTCH connectivity was associated with the outbreak size and time-to-first case, respectively. For each of the three models, we considered two types of covariates: facility-level factors (the degree of connectivity, the number of beds, the proportion of private beds, and for-profit status), and area-level factors (hotspot status). Missing data in the degree of connectivity during our study period was imputed using a missingness model as a function of the baseline degree of connectivity before the pandemic (from 2019-01-01-2020-02-25), assuming missing at random. Given the small sample size, the results are reported using 90% credible intervals (CrI). The detailed model structures are summarized in *Supplementary text* .

Each of the models was calibrated using using8 chains and 10,000 iterations with the *rstan* package (39). All analyses were performed under R version 4.3.1.

### Ethical Approval

Ethics approval was obtained from the Unity Health Toronto Research Ethics Board (#23-198).

## Results

Of the 179 LTCH in Greater Toronto Area, Ontario, 106 (59%) experienced an outbreak (at least one case among residents during the study period, **Table 1** ). Among LTCH with an outbreak, the median cumulative diagnosis rate of SARS-CoV-2 during the study period was 237 per 1,000 residents (interquartile range [IQR]: 4 to 474). The median time-to-first case since the beginning of the epidemic in Ontario (2020-02-26) was 47 (IQR: 33-69) days. LTCH that experienced an outbreak were homes with a larger number of residents, a larger number of beds, and homes with a smaller proportion of beds that were private. A higher proportion of LTCH with outbreaks were located in SARS-CoV-2 neighbourhood hotspots.

**Table 1.**
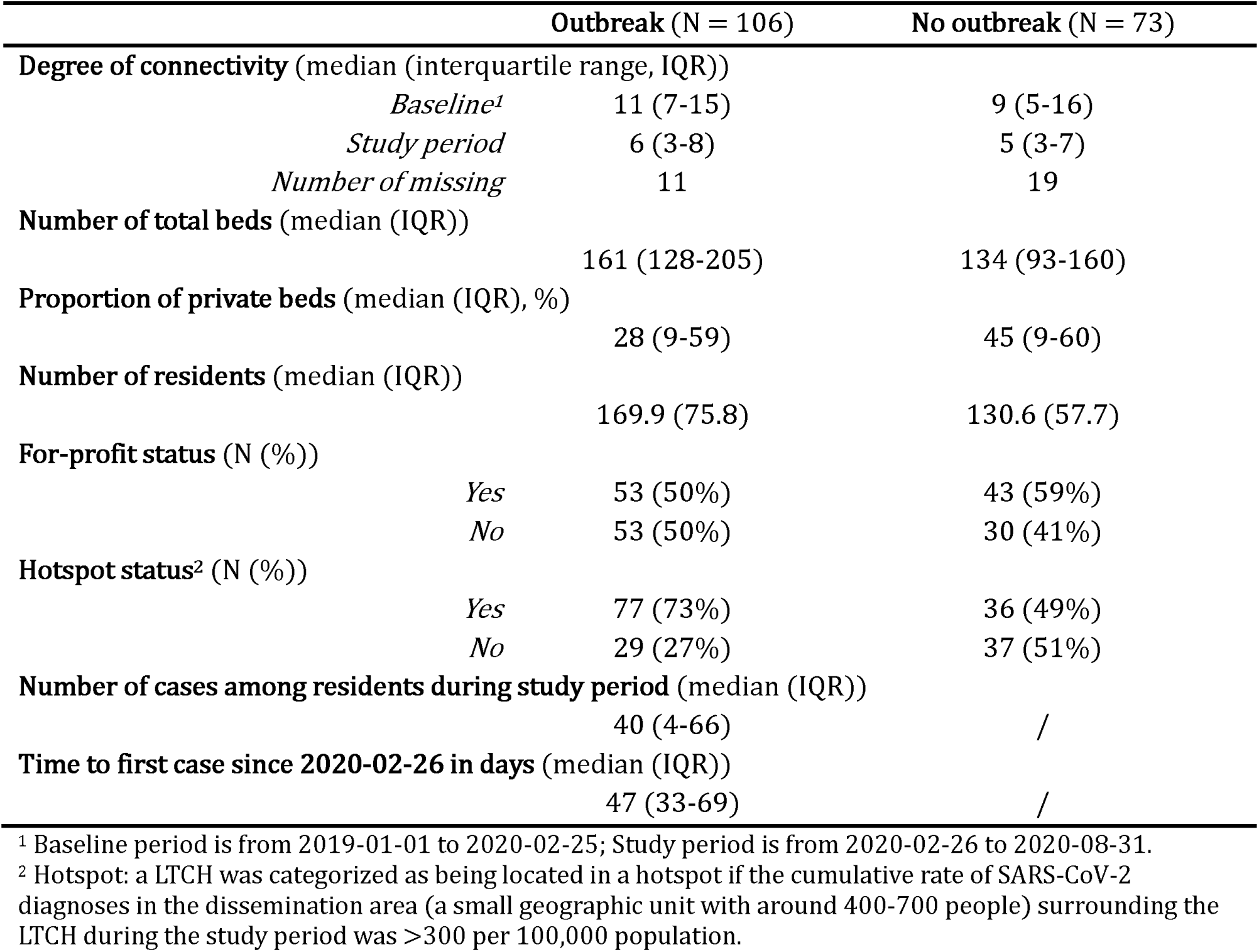
Characteristics of long-term care homes by outbreak status in Greater Toronto Area, Ontario.

The average degree of inter-LTCH connectivity between the LTCH during the study period decreased by 53%, compared to their baseline level. LTCH with an outbreak had slightly higher degree connectivity than those without an outbreak (**Figure 1** ). Regardless of outbreak status, LTCH located in neighbourhood hotspots, with larger number of beds and residents, and with for-profit ownership had higher inter-LTCH connectivity with other LTCH (**Figure 1** ; **Figure S1-4** ). No differences in the degree of inter-LTCH connectivity were observed by proportion of private beds (**Figure S5** ).

**Figure 1.**
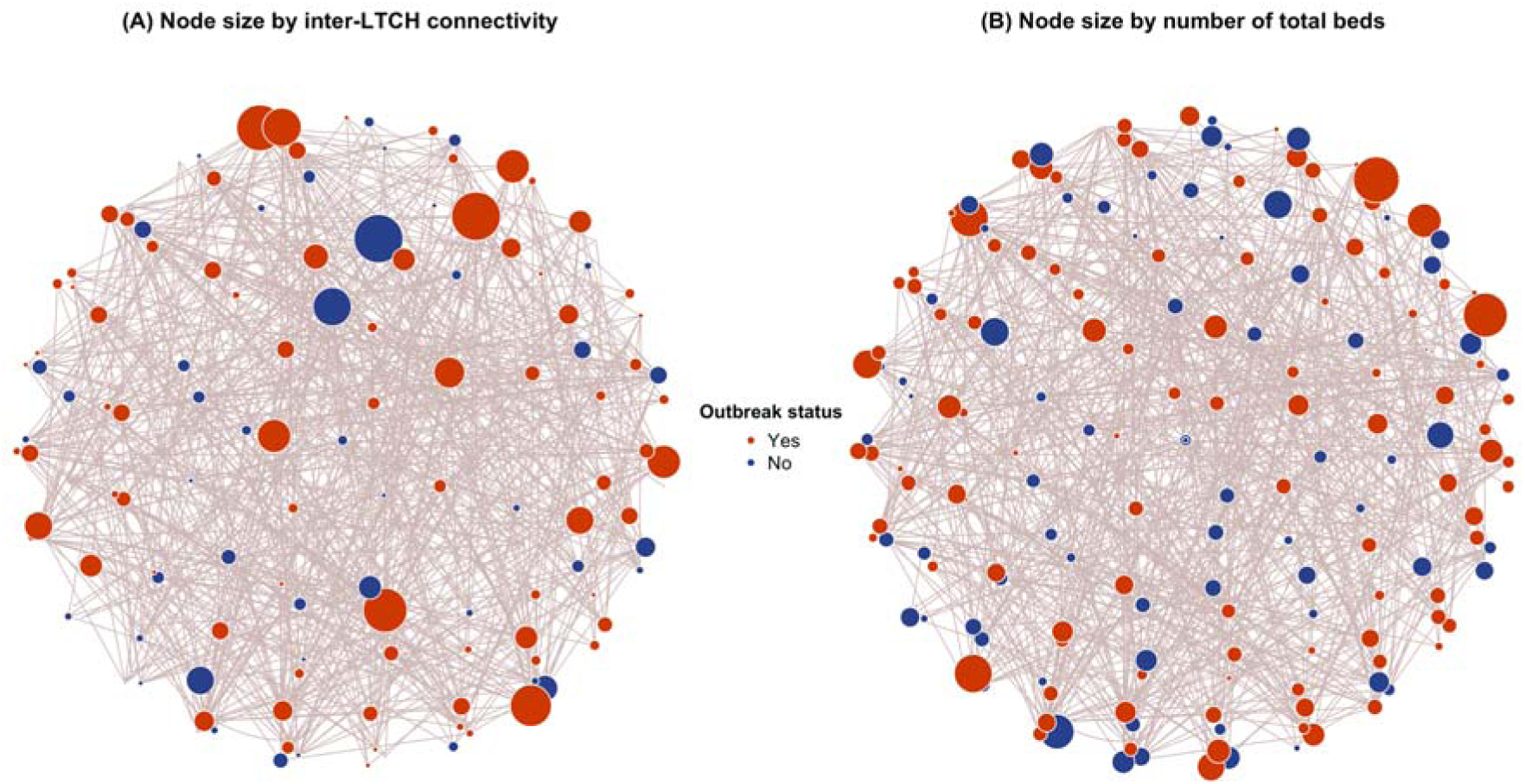
Network connection across long-term care homes (LTCH) in Greater Toronto Area, Ontario by outbreak status. Each node represents a LTCH. The bigger the node, the higher degree of connectivity (panel A) and number of total beds (panel B) that LTCH had between 2020-02-26 and 2020-08-31.

In the unadjusted analysis of our primary outcome, a one-unit increase in inter- LTCH connectivity was associated with an increased odds of a SARS-CoV-2 outbreak (adjusted odds ratio [aOR]=1.11, 90% CrI: 1.03-1.20). The magnitude of association reduced after adjusting for facility-level and neighbourhood-level factors (aOR=1.08, 90% CrI: 1.00-1.17). Analyses of secondary outcomes demonstrated that increased inter-LTCH connectivity was associated with earlier SARS-CoV-2 outbreaks (adjusted hazard ratio [aHR]=1.05, 90%CrI: 1.02-1.09). However, we did not observe a statistically significant association between the degree of connectivity and the outbreak size (adjusted rate ratio [aRR] = 1.04, 90%CrI: 0.96-1.13).

Location of the home in a neighbourhood hotspot was also associated with greater odds of an outbreak (aOR=2.00, 90%CrI: 1.14-3.58) and with the hazard of an earlier first SARS-CoV-2 case (HR = 1.66, 90%CrI: 1.14-2.42). A higher proportion of private beds was associated with a lower odd of an outbreak (aOR=0.28, 90%CrI: 0.08-0.95), a lower hazard for earlier first SARS-CoV-2 cases (aHR=0.47, 90%CrI: 0.24-0.94), and a smaller outbreak size (aRR=0.19, 90%CrI: 0.05-0.80. We did not observe an association of number of total beds and for-profit ownership on all of the three outcomes examined.

## Discussion

Using connectivity information derived from the mobile-device data, we quantified the strength of association between inter-LTCH connectivity and risk, timing, and size an outbreak of SARS-CoV-2 during the first wave of the COVID-19 epidemic. We found that for each additional facility to which an LTCH was connected, there was an 8 percent-point increase in the odds of an outbreak and a 5 percentage-point higher risk of an earlier outbreak. However, there was no evidence of an association between inter-LTHCH connectivity and outbreak size. Facility-level attributes including proportion of private beds and the incidence of SARS-CoV-2 cases in surrounding communities were also strongly associated with the probability and timing of an outbreak in the LTCH.

Our study suggests that inter-LTCH connectivity increased the risk of an outbreak and earlier timing of an outbreak with an exponential augmentation. For example, connections with 10 additional facilities meant 2.2-fold higher odds of an outbreak.

However, connectivity was not associated with the size of an outbreak. Taken together, this association of the first two outcomes suggests that connectivity may represent an important risk factor for the introduction of a pathogen; nevertheless, it becomes less important once nosocomial transmission within a facility takes hold. This interpretation aligns with epidemic theory that staff who worked in multiple facilities posed a higher risk of case importation into these facilities (40). Empirical studies using mobile-device data to measure connectivity in the United States and Canada also found that cross-facility staff movements accounted for approximately 50% of the cases in LTCH (34) and reduced mobility between facilities was associated with lower risk of SARS-CoV-2 in LTCH (41).

Moreover, case studies that explicitly measured staff movement in the United Kingdom provided further evidence of staff-to-resident transmission and elevated risk of infection among staff working at multiple facilities (42). Therefore, these empirical associations (and which were conceptualized as risk factors), alongside epidemic theory, suggest that efforts to reduce connectivity could be an important early strategy but alone would be insufficient to reduce nosocomial transmission risks.

Our findings on other facility-level and area-level factors beyond connectivity align with previous literature on the importance of the facility and community characteristics. These studies demonstrated the important role of lower staff-to-resident ratio, outbreaks in surrounding community, for-profit status, and other architectural structure (e.g., proportion of private beds) factors on risks of SARS-CoV-2 outbreaks in congregated living settings (23, 34, 37, 43, 44). This is expected because more cases in surrounding community can increase the risk of case importation, and lacking space for distancing and isolation increases the risk of transmission among residents. Low staff-to-resident ratio not only decreases the quality of care, but also increases the likelihood of staff not taking sick leave, and therefore increasing the risk of transmission (45). We also found a protective association between the proportion of private beds and the timing of an outbreak which has not yet been discussed elsewhere. A possible explanation could be that LTCH with higher proportion of private beds were newer (better design) and had higher quality of care (46). These findings provide further evidence to support structural strategies within LTCH to reduce transmission risks for SARS-CoV-2, and other respiratory viruses (23, 27, 43).

Our study should be interpreted considering certain limitations. First, we used mobile device connectivity as proxy to staff movement. The quality of the data highly depends on the extent of measurement error and the degree of completeness of the data. This may induce bias if there exists systematic error (e.g., mobile device sensor quality, device signal not captured) and missingness of staff mobile connectivity not at random (e.g., geolocation failure, invalid geocoordinate) (47). For example, if missingness among LTCH with an outbreak was higher than those without an outbreak, then it might bias the effect of connectivity towards the null, and vice versa. However, mobile data is one of the most feasible sources of mobility information for large-scale studies on movement, compared to methods that need to distribute specific devices such as portable sensors. Second, the availability and quality of time-series network connection data were limited. Therefore, we only used overall degree of connectivity as measures of the inter-LTCH movement, instead of also looking into the impact of other network connectivity measures (e.g., the strength of connection, which also takes into account of the total number of visits between linked facility (34)) and the changing connectivity across time. There variables may help generate more precise estimates of the association, especially for the time-to-event analysis.

Moreover, we did not study the mixing pattern of the connections between LTCH due to the quality of the data. For example, staff working at a facility with an outbreak may be more likely to have another job at the other facility, or the linkages were random, or those who moved between facilities were more likely to be personal supporting staff who had direct contact with residents (48). This information can help estimate the causal relationship between the staff movement and the outbreaks in LTCH. Future efforts should be paid to overcome these data limitations.

This study also has several strengths. First, we used mobile data to measure the mobility of LTCH workers. This type of data provides valid and nearly real-time population movement information at a large scale that can hardly be achieved by traditional surveys (49, 50). Moreover, this study adopted a Bayesian approach to deal with the missing data, which incorporated the uncertainty directly into the model and provided more robust and realistic estimates.

These findings have two key implications. First, if most of the connectivity between facilities is due to movement of staff who work in multiple facilities, then our findings suggest that implementing a “work only in one-site” strategy (staff cohorting) could help reduce risks of an outbreak but may not reduce the size of an outbreak. The latter is critical given trade-offs that have been identified and include low staffing levels associated with higher rates of SARS-CoV-2 in LTCH (4), staff burnout (24), lower care quality and pressure on LTCH residents (25), etc. A pandemic response strategy that carefully separates strategies to reduce “importation” risks and “nosocomial” risks, therefore, can balance these trade-offs. For example, structural strategies to reduce nosocomial transmission risks, such as ensuring training of LTCH staff and strict protocols of infection control, increasing staff-residence ratio and adequate staff sick pay, and improving living environment of the residents (17, 23, 27), are particularly important because it is the nosocomial risks and outbreak size that shape the burden of morbidity and mortality. Second, our findings on the importance of location (in a hotspot) highlight the critical role that reducing transmission in the community could have on reducing importation risks in facilities (43).

## Conclusion

To prevent and manage outbreaks of SARS-CoV-2 and other respiratory viruses in long-term care homes, policies should focus on limiting staff movement between facilities to reduce virus importation. Simultaneously, addressing widespread staffing shortages is critical to ensure care quality without compromising infection control. Structural strategies, such as improving infection prevention protocols and facility infrastructure, are essential to minimize nosocomial transmission and enhance outbreak preparedness in these congregate living settings.

## Data Availability

All data except for long-term care home SARS-CoV-2 cases and occupation are publicly available information.

## Acknowledgements

We thank Kristy Yiu (Unity Health Toronto) and Mackenzie Hamilton (Unity Health Toronto) for data management support.

## Author contributions

YX, HM and SM conceived of and designed the study. YX conducted the literature search, developed and conducted the statistical analysis, and drafted the manuscript with editing by SM. KM, QH, SA, JMB, IF, JJ, KK, JM, and GM supported data curation. HM, KM, SES, CF, GM, QG, SA, JMB, IF, JJ, KK, JM, LW, AKC, SDB, MMG, and SM interpreted results, critically reviewed and edited the article.

