## Supplementary figures for "Connectivity between long-term care homes and subsequent SARS-CoV-2 outbreaks"

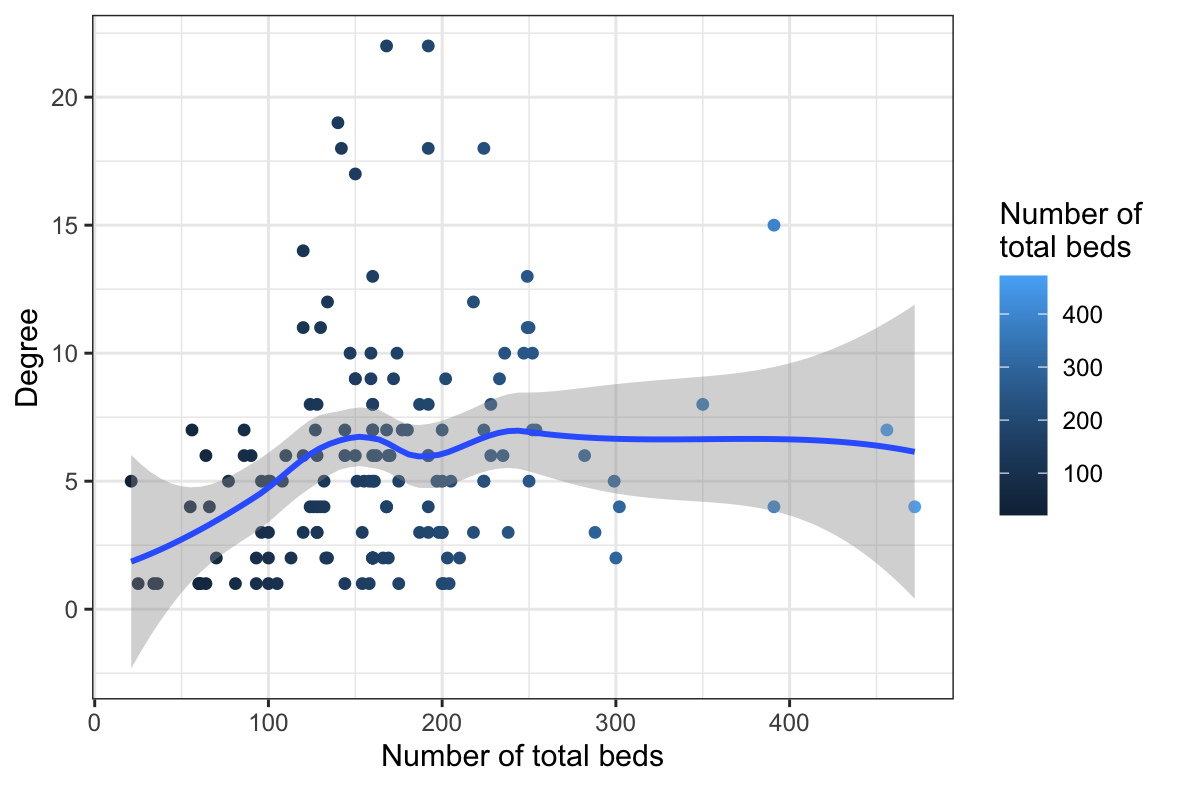

**Figure S1**. Degree of connectivity of long-term care homes in Greater Toronto Area, Ontario by number of total beds.

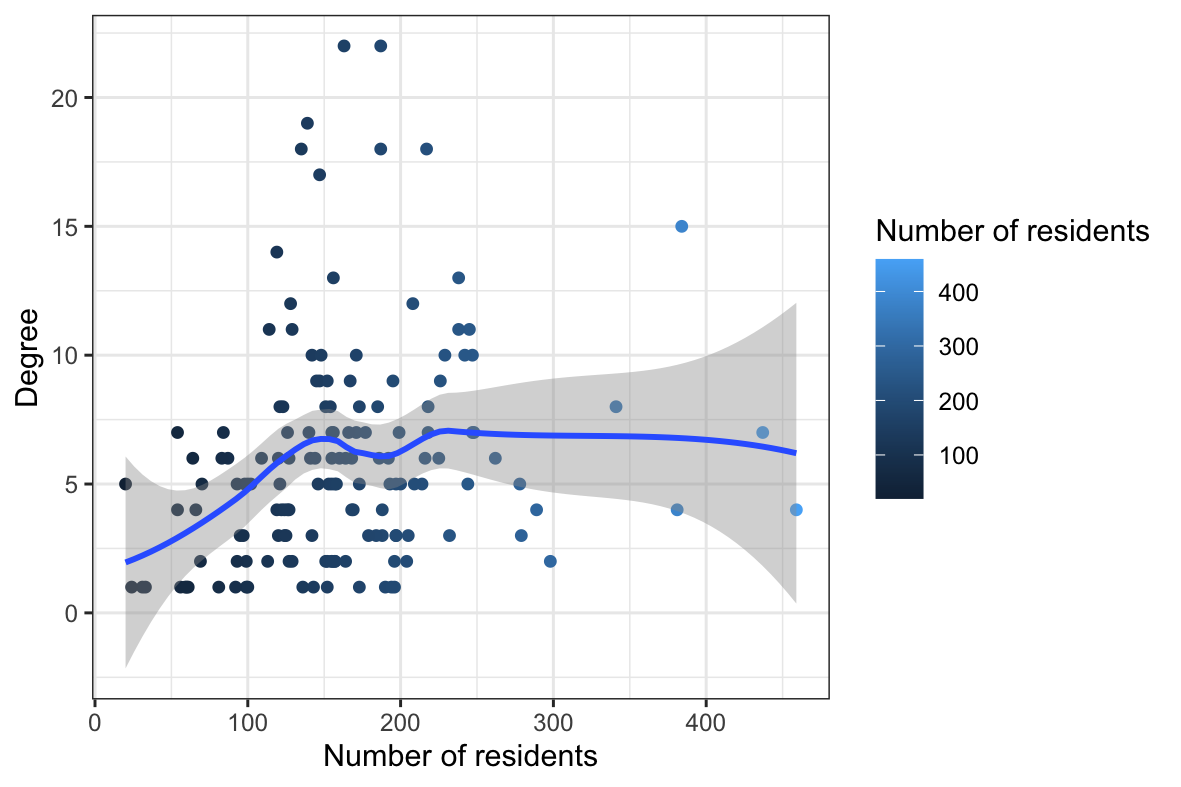

**Figure S2**. Degree of connectivity of long-term care homes in Greater Toronto Area, Ontario by number of residents.

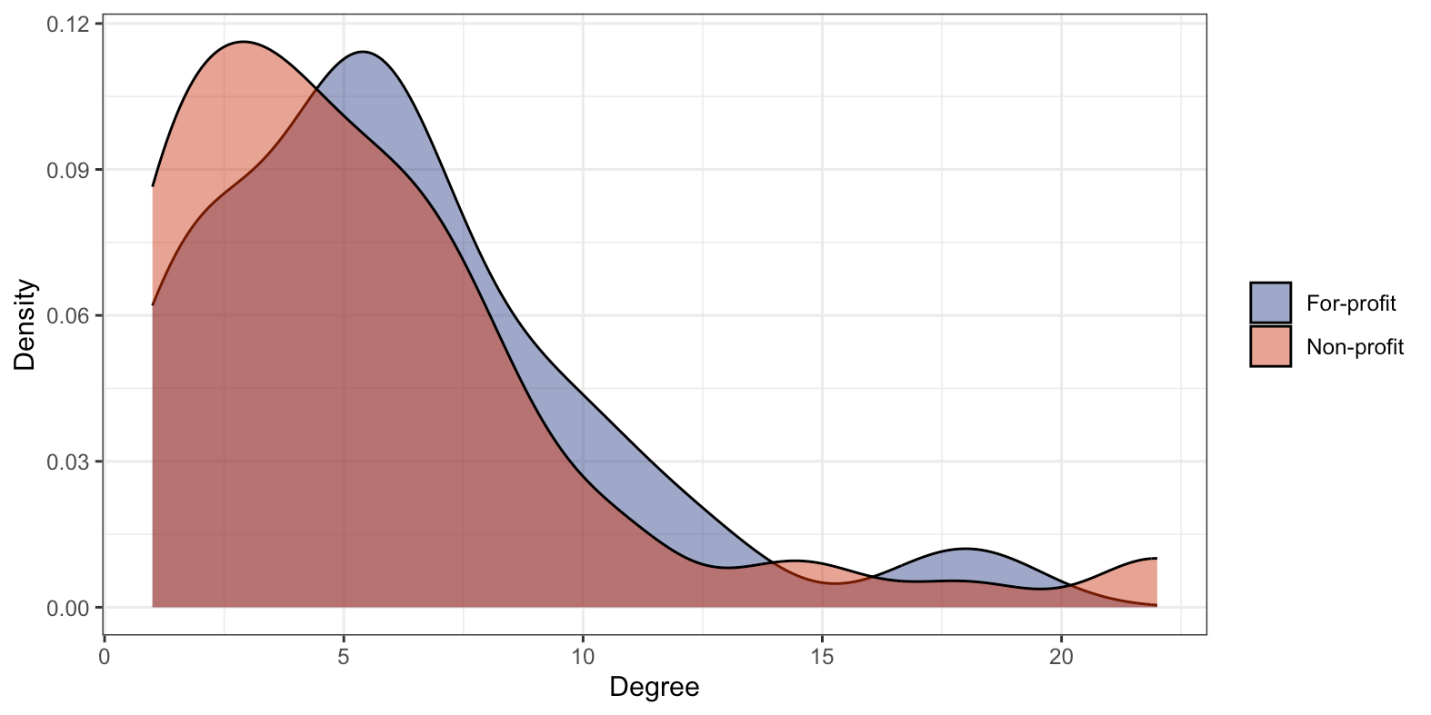

**Figure S3**. Degree of connectivity of long-term care homes in Greater Toronto Area, Ontario by for-profit status.

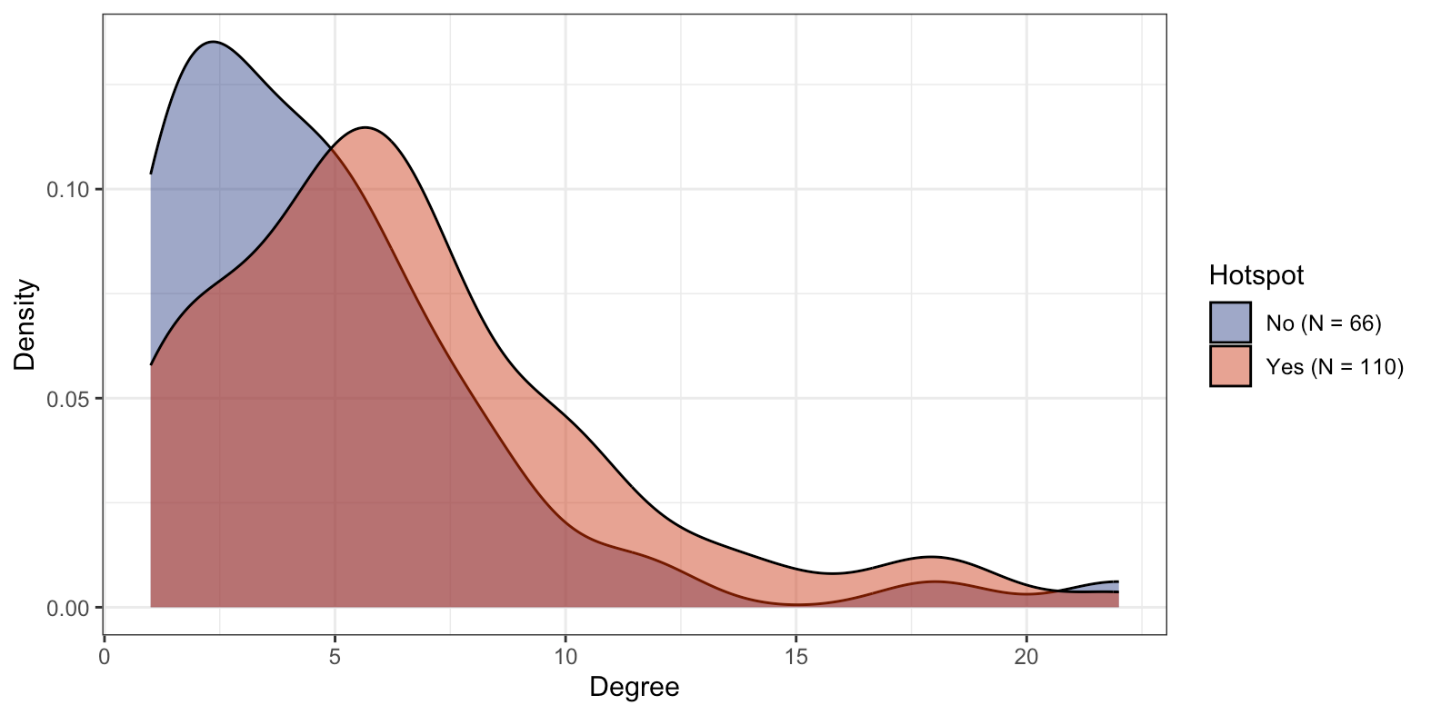
**Figure S4**. Degree of connectivity of long-term care homes in Greater Toronto Area, Ontario by hotspot status.

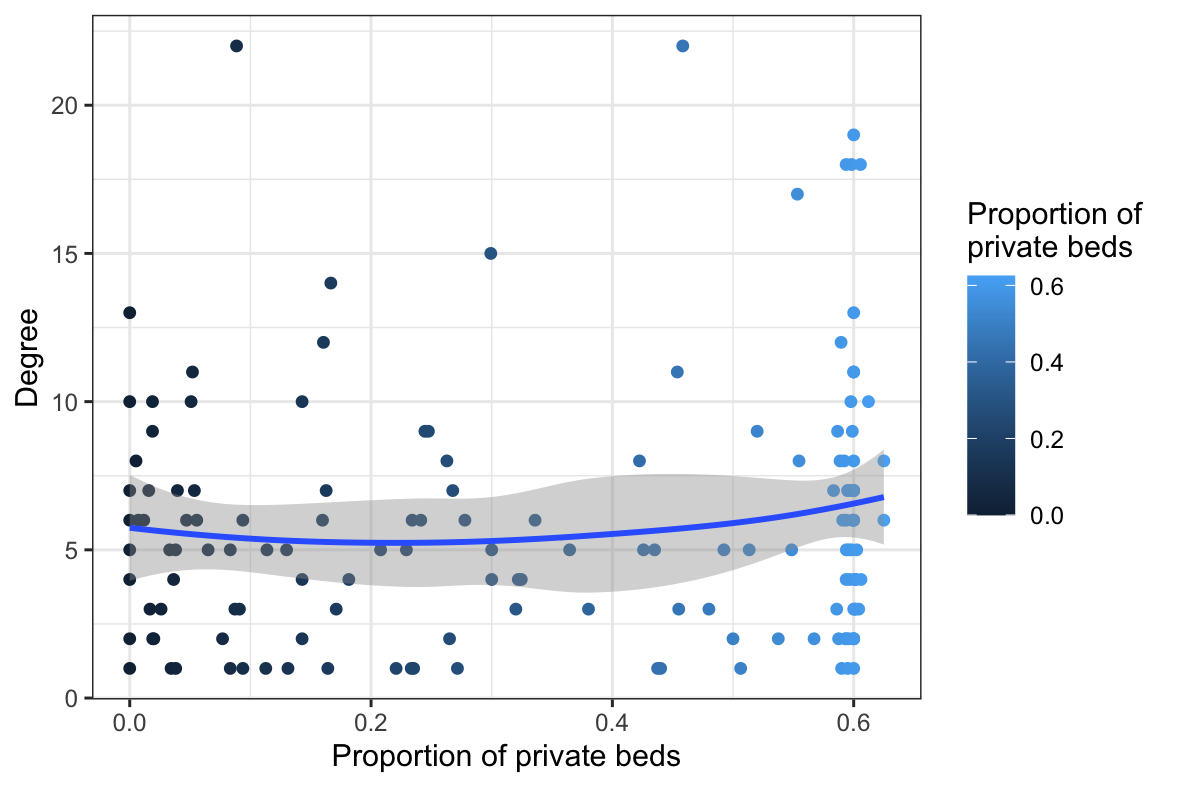

**Figure S5**. Degree of connectivity of long-term care homes in Greater Toronto Area, Ontario by proportion of private beds.
