## Supplementary text for "Connectivity between long-term care homes and subsequent SARS-CoV-2 outbreaks"

Model 1

$$Oubtreak_{i} \sim\boldsymbol{Bernoulli}(p_{i})$$

$$logit(p_{i})=\beta_{0}+\beta_{1}Degree_{i}+\beta_{2}N_{bed_{i}}+\beta_{3}Prop_{private_{i}}+\beta_{4}Profit_{i}+\beta_{5}Hotspot_{i}+\epsilon_{i}$$

Model 2

$$Total case_{i} \sim\boldsymbol{NegBin}\left( \lambda_{i}, \alpha\right)$$

$$\log\left( \lambda_{i} \right)=\beta_{0}+\beta_{1}Degree_{i}+\beta_{2}N_{bed_{i}}+\beta_{3}Prop_{private_{i}}+\beta_{4}Profit_{i}+\beta_{5}Hotspot_{i}+\beta_{6}N_{resident_{i}}+\epsilon_{i}$$

Model 3

$$h(t\mid\boldsymbol{x}i)=\lambda_{0}(t)exp(\beta_{0}+\beta_{1}Degree_{i}+\beta_{2}N_{bed_{i}}+\beta_{3}Prop_{private_{i}}+\beta_{4}Profit_{i}+\beta_{5}Hotspot_{i}+\epsilon_{i})$$

where i represents a single long-term care facility; the dispersion parameter $\alpha\sim N(0, \sigma_{\alpha}^{2}); \sigma_{\alpha} \sim Gamma(0.1, 0.1)$; $h(t\mid\boldsymbol{x}_{i})$ represents the hazard function given all covariates; and $\lambda_{0}(t)$ represents the baseline hazard function. Prior distributions of $\boldsymbol{N}\left( 0, 100 \right)$ were assigned to all coefficient parameters ($\beta_{0-6}$).

To handle missing data in the degree of connectivity during our study period, the following missingness model was assumed under the assumption of missing at random.

$$Degree_{i} \sim\boldsymbol{N}(\varphi_{1}Baseline Degree_{i} + \varphi_{2}Outbreak_{i}, \sigma_{Degree}^{2})$$

where $\varphi_{1,2} \sim\boldsymbol{N}\left( 0, 100 \right)$ and $\sigma_{degree} \sim Gamma(0.1, 0.1)$.
